# Natural Language Processing for assessing multimorbidity: A systematic review

**DOI:** 10.1101/2025.06.30.25330608

**Authors:** Ravi Shankar, Ziyu Goh, Xu Qian

## Abstract

**Background:** Multimorbidity poses significant healthcare challenges globally. Current assessment methods rely primarily on structured electronic health record (EHR) data, potentially missing valuable information contained in unstructured clinical notes. Natural language processing (NLP) techniques offer promising solutions for extracting comprehensive multimorbidity data from these unstructured sources.

**Objectives:** To identify, characterize, and critically appraise studies utilizing NLP techniques for multimorbidity assessment from unstructured EHR data.

**Methods:** This systematic review will follow PRISMA-P guidelines and be conducted according to Cochrane and Joanna Briggs Institute methodologies. We will search multiple databases (PubMed, Web of Science, Embase, CINAHL, MEDLINE, Cochrane Library, PsycINFO, and Scopus) from inception to February 2025. Eligible studies will include adult populations with multimorbidity (≥2 chronic conditions) where NLP techniques were applied to unstructured EHR data and compared against reference standards. Two independent reviewers will screen studies, extract data, and assess methodological quality using PROBAST, CLAIM, TRIPOD, and QUADAS-2 tools as appropriate. Both quantitative synthesis (meta-analysis) and narrative synthesis will be considered based on study heterogeneity.

**Outcomes:** Primary outcomes include validity metrics of NLP-based multimorbidity assessment (sensitivity, specificity, positive predictive value, F1 score, AUROC). Secondary outcomes include reliability measures, generalizability assessments, efficiency metrics, and end-user perspectives.

**Discussion:** This review will establish the current state of evidence on NLP for multimorbidity assessment, identify best practices and challenges, and guide future research efforts in this emerging field. Findings will inform the development and implementation of improved methods for extracting multimorbidity information from unstructured clinical text, potentially enhancing risk stratification, care planning, and research for patients with multiple chronic conditions.

## Background

Multimorbidity, defined as the presence of two or more chronic conditions in an individual, represents a growing global healthcare challenge driven by population aging, increased life expectancy, and shared risk factors across conditions (Johnston et al., 2019; McMahon et al., 2018; Xu et al., 2017). Patients with multimorbidity face heightened risks of functional decline, poor quality of life, care fragmentation, polypharmacy, and premature mortality (Fortin et al.,2004; Nunes et al, 2016; Violan et al., 2014). While accurate measurement of multimorbidity is foundational for risk stratification, care planning, quality improvement, and research (Nicholson et al., 2019), current assessment approaches vary widely in methods and data sources, limiting comparability and generalisability of findings (Holzer et al., 2017; Willadsen et al., 2016).

Furthermore, current data collection relies heavily on labour-intensive patient self-reports or interviews, and on medical records and administrative databases (Ho et al., 2021), which may not fully capture the spectrum and granularity of health conditions, particularly those not consistently coded (Endut et al., 2022).

Unstructured clinical notes contain valuable detailed information about patients’ medical history that complements structured EHR data (Ford et al., 2016). However, manual extraction from free-text is impractical at scale. Natural language processing (NLP) offers a promising solution for efficiently mining multimorbidity data from unstructured text (Koleck et al, 2019), automatically identifying clinical concepts, mapping them to standardized terminologies, and inferring relationships between concepts (Luo et al., 2017; Sheikhalishahi et al., 2019).

In multimorbidity contexts, NLP can provide more comprehensive understanding of disease burden and interactions than structured data alone. By processing large volumes of clinical notes, NLP can identify co-occurring conditions often missed in structured fields, characterize multimorbidity patterns over time, and uncover complex relationships between conditions (Wang et al., 2018). However, applying NLP to multimorbidity assessment remains an emerging area with unique challenges warranting further investigation.

### Policy relevance

According to the World Health Organisation (WHO) in 2020, healthy ageing has replaced their focus on active ageing. The core concepts are diversity – that all the elderly cannot be lumped into the same group – and inequity – that the accumulation of experiences over one’s life result in the situation we observe the elderly in.

The integration of NLP techniques for multimorbidity assessment supports recent healthcare policy trends toward data-driven, personalized care models and value-based healthcare that consider the complex interplay of multiple conditions (Muth et al., 2019). Improved multimorbidity assessment through NLP could mean more efficient resource allocation and enhanced disease surveillance, thereby supporting public health policy objectives for chronic disease management. NLP can enable more granular phenotyping of multimorbidity by extracting rich contextual information such as disease severity, temporality, and attribution (Spasic & Nenadic, 2020).

### Objectives

1. What is the range of NLP methods, tools, and data sources used for multimorbidity assessment, including pre-processing techniques, annotation schemes, modeling architectures, and clinical targets? How do they differ in terms of data requirements, clinical targets, and performance?
2. How well do NLP approaches perform for identifying and characterizing multimorbidity compared to reference standards, using metrics such as sensitivity, specificity, positive predictive value, and F1 score?
3. What are the main challenges and limitations of using NLP for multimorbidity assessment, and how can these be addressed through improved methods and implementation strategies?
4. What are factors associated with better or worse performance of NLP methods, such as clinical setting, patient population, multimorbidity definitions, and gold standards used for training and evaluation?
5. What is the methodological quality of included studies and potential risk of bias using established tools and frameworks?
6. What is the overall state of the evidence? What are the key findings, limitations, and areas for future research and implementation?

Why is this review needed in light of existing reviews?

Despite the growing recognition of NLP as a valuable tool for extracting clinical insights from unstructured EHR data, there has been no systematic effort to synthesize the evidence on its use specifically for multimorbidity assessment. Prior reviews have examined the application of NLP for a range of clinical tasks, such as drug safety surveillance (Wu et al., 2019), clinical trial eligibility screening (Chapman et al., 2011), and quality measure reporting (Shivade et al., 2014), but none have focused on multimorbidity as a distinct use case with its own set of considerations and challenges. A systematic review is needed to establish the current state of the science, identify best practices and common pitfalls, and guide future research and development efforts in this important area.

Campbell, Cochrane, INPLASY, and the Registry of Systematic Reviews/Meta-Analyses in Research Registry were searched. There returned no results of a combination of the words “natural language processing” and “multimorbidity” or “multiple chronic conditions”.

This protocol has been registered in PROSPERO (Registration CRD420251007223).

### Inclusion criteria

The protocol for this systematic review is reported in accordance with the Preferred Reporting Items for Systematic Review and Meta-Analysis Protocols (PRISMA-P) 2015 statement (Moher et al., 2015). The review will be conducted following methodological guidance from the Cochrane Handbook for Systematic Reviews of Interventions (Chandler et al., 2019) and the Joanna Briggs Institute Manual for Evidence Synthesis (Aromataris & Munn, 2020).

#### Population

The population of interest includes adult patients (aged 18 years and older) with multimorbidity, defined as the presence of two or more chronic conditions. Studies involving only pediatric or adolescent populations will be excluded. No restrictions will be placed on clinical settings, which may include primary care, specialty care, acute care hospitals, and long-term care facilities.

#### Intervention/Exposure

The intervention or exposure of interest is the application of natural language processing techniques to assess multimorbidity from unstructured electronic health record data. This may include rule-based approaches, traditional machine learning algorithms, deep learning neural networks, or hybrid methods. Studies that use NLP only for single disease or condition will be excluded, as the focus is on multimorbidity as a complex construct involving multiple conditions. No diseases or conditions will be excluded. Studies that examine the use of NLP with structured data (e.g., diagnosis codes) will be considered if unstructured data are also analyzed. No specific electronic health records systems or software will be excluded.

#### Comparator

Eligible studies should compare the performance of NLP-based multimorbidity assessment to a reference standard, such as simple counts of diseases, ICD codes, morbidity indices, or other validated measures of multimorbidity. Studies without any comparator will be excluded.

#### Outcomes

The primary outcome of interest is the validity of NLP-based multimorbidity assessment, as measured by metrics such as:

- Sensitivity (recall): The proportion of actual multimorbidity cases that are correctly identified by NLP
- Specificity: The proportion of actual non-multimorbidity cases that are correctly identified by NLP
- Positive predictive value (precision): The proportion of NLP-identified multimorbidity cases that are true positives
- Negative predictive value: The proportion of NLP-identified non-multimorbidity cases that are true negatives
- F1 score: The harmonic mean of precision and recall
- Area under the receiver operating characteristic curve (AUROC): A plot of sensitivity versus 1-specificity across different classification thresholds

Secondary outcomes that may be considered include:

- Reliability of NLP algorithms, such as inter-annotator agreement and intra-annotator agreement
- Generalizability of NLP performance across different patient populations, clinical settings, or institutions
- Efficiency and computational performance of NLP methods
- Qualitative perceptions of end users (e.g. clinicians, researchers) regarding the utility and usability of NLP-based multimorbidity assessment

#### Study design

Eligible study designs will include:

- Development and validation studies of NLP algorithms for multimorbidity assessment
- Comparative studies evaluating the performance of NLP methods against other approaches
- Cohort or case-control studies using NLP to characterize multimorbidity patterns and outcomes
- Qualitative studies exploring stakeholder perspectives on NLP for multimorbidity assessment

The following types of publications will be excluded:

- Commentaries, editorials, and opinion pieces
- Literature reviews and systematic reviews
- Case reports and case series
- Animal studies
- Conference abstracts and posters

#### Information Sources and Search Strategy

A comprehensive literature search will be conducted across multiple electronic databases, including PubMed, Web of Science, Embase, CINAHL, MEDLINE, The Cochrane Library, PsycINFO, and Scopus. These databases have been selected to ensure a broad and rigorous exploration of relevant studies, capturing a wide range of published research across various disciplines.

The initial search will be performed in PubMed and then adapted for the other databases. The search strategies will be developed in consultation with a medical librarian experienced in systematic reviews. A combination of Medical Subject Headings (MeSH) terms and keywords will be used, covering the key concepts of multimorbidity, chronic conditions, electronic health records, natural language processing, and machine learning. The searches will not be limited by language, publication date, or study design. The search will cover the time period from each database’s inception to February 2025. A draft PubMed search strategy is provided below:

(((((“Multimorbidity”[MeSH] OR multimorbid*[tiab] OR “multiple chronic conditions” OR “multiple morbidity” OR “multiple illnesses” OR “multiple diseases” OR polymorbidity OR “complex needs”))) AND (((“Natural Language Processing”[MeSH] OR “natural language processing” OR “medical language processing” OR “clinical language processing” OR “text mining” OR “machine learning” OR “deep learning” OR “neural networks” OR “named entity recognition” OR “concept extraction”))) AND (((“Electronic Health Records”[MeSH] OR “electronic medical record*” OR “electronic patient record*” OR “electronic health record*” OR “computerized patient record*” OR “computerized medical record*” OR “clinical note*” OR “clinical document*” OR “free text” OR “unstructured data”))))

In addition to the electronic database search, the reference lists of included studies and relevant systematic reviews will be hand-searched to identify any additional eligible studies. Grey literature sources such as preprint servers (e.g., medRxiv, bioRxiv), conference proceedings (e.g., AMIA, HIMSS), dissertations and theses databases (e.g., ScholarWorks, ProQuest), and clinical trial registries will also be searched. Finally, experts in the field of clinical NLP and multimorbidity research will be consulted to identify any ongoing or unpublished studies.

### Study Selection

The study selection process will be managed using Covidence systematic review software (www.covidence.org). First, all records identified from the electronic database searches will be imported into Covidence for de-duplication. The unique records will then be screened by two independent reviewers based on the title and abstract. Any discrepancies will be resolved through discussion or adjudication by a third reviewer. The full-text articles of potentially eligible studies will be retrieved and assessed against the pre-defined inclusion and exclusion criteria by the two reviewers independently. Reasons for exclusion at the full-text stage will be recorded and reported. Any disagreements on the final study selection will again be resolved by consensus or a third reviewer. The study selection process will be documented using a PRISMA flow diagram.

### Data Extraction

A standardized data extraction form will be developed in Covidence and piloted on a sample of 5-10 included studies to ensure comprehensive data capture. Two independent reviewers will extract data from each included study using the finalized form, with discrepancies resolved through discussion or third-reviewer arbitration if necessary. The extraction will encompass study characteristics (author, publication year, country, funding); study design (objectives, setting, timeframe, eligibility criteria); sample characteristics (size, demographics, clinical profiles, multimorbidity definitions); NLP methodologies (data sources, preprocessing techniques, annotation methods, modeling approaches); performance metrics (validity, reliability, generalizability assessments); implementation details (computational requirements, processing time, user evaluations); and additional relevant outcomes including challenges and limitations (details in Appendix 1). For missing or unclear information, corresponding authors will be contacted via email, with data marked as “not reported” after three unsuccessful attempts.

### Risk of Bias Assessment

The risk of bias and methodological quality of included studies will be comprehensively assessed using PROBAST+AI (Moons et al., 2025), the updated quality, risk of bias, and applicability assessment tool for prediction models using regression or artificial intelligence methods. For NLP model development and validation studies, we will apply the Prediction model Risk Of Bias ASsessment Tool-Artificial Intelligence (PROBAST+AI). PROBAST+AI consists of two distinctive parts: model development and model evaluation. For model development, PROBAST+AI users assess quality and applicability using 16 targeted signaling questions. For model evaluation, PROBAST+AI users assess the risk of bias and applicability using 18 targeted signaling questions. Both parts contain four domains: participants and data sources, predictors, outcome, and analysis. Each domain contains signaling questions to guide the assessment, and the overall judgment for each domain is rated as low, high, or unclear risk of bias.

PROBAST+AI has been specifically designed to address all novel and necessary methodological considerations for a broader set of modelling approaches than only prevailing statistical techniques, making it suitable for assessing NLP-based prediction models for multimorbidity. We will also evaluate adherence to the TRIPOD+AI statement, which provides updated guidance for reporting clinical prediction models that use regression or machine learning methods. Adherence to TRIPOD+AI will be assessed using a published adherence assessment form, which evaluates the completeness of reporting across the TRIPOD+AI main items and sub-items. The proportion of items reported will be calculated and summarized descriptively.

For studies evaluating the performance of NLP algorithms in identifying multimorbidity, we will focus on the model evaluation component of PROBAST+AI, which assesses the risk of bias and applicability in the evaluation of prediction model performance. Two reviewers will independently assess quality, risk of bias, and applicability for each included study using PROBAST+AI. Disagreements will be resolved by consensus or arbitration by a third reviewer. The quality and risk of bias judgments for each domain and overall will be presented in summary tables and figures, following the PROBAST+AI guidance. Adherence to TRIPOD+AI will be reported as the proportion of items fully, partially, or not reported.

The risk of bias and reporting quality assessments will inform the interpretation and discussion of review findings. Studies with high risk of bias or poor reporting will be identified and their limitations explicitly considered when formulating conclusions and recommendations. However, studies will not be excluded based on risk of bias or reporting quality, as the aim is to comprehensively map the current state of the field and identify areas for improvement. Sensitivity analyses may be conducted to explore the impact of risk of bias on pooled performance estimates if meta-analysis is performed.

### Data Synthesis

The approach to evidence synthesis will depend on the nature and heterogeneity of the included studies. If enough studies (e.g., ≥10) with comparable NLP methods, multimorbidity measures, and outcome metrics are available, a quantitative meta-analysis of performance measures (e.g., sensitivity, specificity, F1 score) will be considered. A random effects model would be used to account for both within- and between-study variability. Forest plots will be generated to visualize the pooled estimates and confidence intervals. Statistical heterogeneity will be assessed using the I2 statistic and Cochran’s Q test. Potential publication bias will be examined using funnel plots and Egger’s regression test.

If formal meta-analysis is not appropriate due to clinical, methodological, or statistical heterogeneity, a narrative synthesis of the evidence will be conducted following the Synthesis Without Meta-analysis (SWiM) guideline (Heus et al., 2019). The narrative synthesis will focus on describing patterns, similarities, and differences across studies in terms of NLP methods, multimorbidity definitions, data sources, performance metrics, and implementation considerations. Tables and figures will be used to summarize key study characteristics and findings. Subgroup analyses may be performed to explore potential effect modifiers, such as clinical setting, disease types, and algorithm architectures. The strength and quality of the evidence will be assessed using the Grading of Recommendations Assessment, Development and Evaluation (GRADE) approach (Guyatt et al., 2008).

## Data Availability

All data produced in the present work are contained in the manuscript

## Roles and responsibilities

The review team consists of two researchers with complementary expertise. One member brings content expertise in multimorbidity through comprehensive literature review and ongoing research in this field. The second member contributes extensive methodological rigor as an experienced systematic reviewer with strong statistical analysis skills. Both members will collaborate on information retrieval, with consultation from the university’s medical librarian as needed to ensure comprehensive search strategies.

